# Loss of regulation of complement C5 activation in HIV associated Preeclampsia

**DOI:** 10.1101/2022.07.06.22277325

**Authors:** Sumeshree Govender, Takafira Mduluza, Louansha Nandlal, Thajasvarie Naicker

## Abstract

**Objective:** Maternal mortality remains a global health concern in developing countries that are also affected by HIV infection. Complement components are anaphylatoxin that mediate several growth factors necessary during pregnancy. An extensive stimulation of the complement system contributes to the pathogenesis of preeclampsia; hence its inhibition facilitates a successful pregnancy. The study evaluated the expression of complement components C2 and C5a in HIV and the association with preeclampsia.

**Materials and Methods:** Serum samples were collected from 76 pregnant women of which 38 were preeclamptic and 38 normotensive pregnant. The participants were further stratified according to HIV infection status. Bio-Plex multiplex immunoassay method was used to quantify serum concentration of C5a and C2 complement components.

**Results:** The C2 complement concentration was not significantly different between preeclamptic and normotensive pregnant women, irrespective of HIV status as well as pregnancy type. However, based on preeclamptic *vs* normotensive pregnancy type, the expression of C5a was significantly different (*p* = 0.05). The C5a levels were downregulated in preeclampsia compared to normotensive women, irrespective of HIV status. Both C2 and C5a concentrations did not differ across all study groups.

**Conclusion:** This novel study reports a loss of regulation of complement activation shown by the downregulation of C5a in preeclamptic compared to normotensive pregnant women, regardless of HIV status. Complement dysregulation affects the host innate defence, and as a consequence, intensifies placental and fetal injury. Moreover, HIV status did not influence the expression of both C5a and C2, irrespective of pregnancy type, this may be attributed to Highly Active Antiretroviral Therapy.

## Introduction

Maternal mortality remains a global health concern particularly in low- and middle-income countries [1]. While Human Immunodeficiency Virus (HIV) infection, haemorrhage, and hypertensive disorders in pregnancy (HDP) are the main factors contributing to maternal mortality in South Africa [2]. Preeclampsia (PE) is a common and potentially fatal HDP, affecting 3-8% of all pregnancies worldwide [3]. It is diagnosed by a new onset high blood pressure of ≥140/90 mmHg with/without proteinuria (> 300 mg/d) and/or evidence of maternal organ dysfunction at or after 20 weeks of gestation [4]. PE is a two-stage disorder in which stage 1 involves abnormal placentation shown by inadequate trophoblast invasion with resultant deficient myometrial spiral artery remodelling that leads to an ischemia/ hypoxic micro-environment [5]. The maternal syndrome of PE is a consequence of ischaemia which leads to widespread endothelial damage, multi-organ involvement, and the clinical signs and symptoms of PE or stage 2 of this pregnancy disorder [6].

During normal pregnancy, the adaptations of the innate and adaptive immune system ensure the survival of the fetus [7]. However, during PE, immune hyper-reactivity results in maternal intolerance of the fetus [8]. In contrast, in HIV infected individuals, the immune response is suppressed, thus in HIV associated PE a neutralisation of immune response is expected [9, 10]. However, the use of Antiretroviral Therapy (ART) causes reconstitution of the immune system [11], thereby predisposing HIV infected women to the development of PE [12]. The complement cascade, a fundamental part of the innate immune system is enhanced in the pathogenesis of PE [13-15], hence its inhibition facilitates a successful pregnancy [16]. Notably, the complement system is activated via one of three pathways *viz*., classical, lectin, or alternative [17] that merge at a central point of C3 convertase production [18]. Thereafter cleavage of C3 convertase leads to a central terminal path that mediates host defence via the opsonization of pathogens, eliciting inflammation, lysis of pathogen cells, and/or clearing of immune complexes [15, 17, 19-21]. However, the role of the complement system in HIV infection is complex, as it may protect the host from HIV infection whilst also augmenting the infectivity of HIV [22].

Anaphylatoxins are potent inflammatory mediators targeting a vast spectrum of immune and non-immune cells [23]. Complement component 5a (C5a) is a powerful anaphylatoxin [15, 24, 25]. C5a mediates the release of potent anti-angiogenic factors, soluble vascular endothelial growth factor receptor 1 (sFlt-1) and soluble endoglin (sEng) [26] with a concomitant decline in pro-angiogenic growth factors including placental growth factor (PIGF) and vascular endothelial growth factor (VEGF) [26, 27]. Additionally, increased C5a elevates pro-inflammatory cytokines such as Tumour Necrosis Factor-alpha (TNF-α) and Interleukin 6 (IL-6) both known to promote HIV infection [25, 28].

Complement component 2 (C2) is a precursor protein and is a critical factor in both the classical pathway (CP) and lectin pathway (LP) [29, 30]. C2 is cleaved to form C3 convertase, C4bC2a [6, 24]. The exact role of C2 in pregnancy and during HIV infection remains unknown. Nonetheless, existing research shows that C2 concentrations are high in pregnancy [16]. Moreover, complement components C1q – C4 are increased in asymptomatic HIV-infected patients compared to healthy controls [31]. In light of the paucity of information available on the regulation of complement components in HIV associated PE, this study aimed to evaluate the concentration of serum C5a and C2 during pregnancy.

## Materials and Methods

### Ethical Approval

This prospective study utilized stored serum samples for which institutional ethics class approval (BCA 338/17) was obtained. Written informed patient consent, hospital manager’s approval and the regulatory authority consent were obtained in the primary study. Participants confidentiality was respected, and participant details remained anonymous when processing the samples.

### Study population

Post consultation with an institutional biostatistician, sample size was calculated using G*Power statistical software (80% power and 95% probability). To detect a moderate effect size of 0.66 between two groups normotensive *vs* preeclamptic women or HIV positive *vs* HIV negative assuming equal groups (n=38 per group), a sample size of 76 pregnant women was required. To compare four groups, normotensive (HIV+ and HIV-) and preeclamptic (HIV+ and HIV-), a sample size of 19 in each group was needed to detect a large effect size of 0.95. A study population of 76 was recruited from a large regional hospital, consisting of 38 normotensive and 38 preeclamptic women. Both groups were further stratified by HIV status into 19 HIV positive preeclamptic and 19 HIV non infected preeclamptic, also 19 HIV positive normotensive pregnancy and 19 HIV non-infected normotensive pregnant women.

#### Inclusion Criteria

the study group consisted of primigravid and multigravida participants, diagnosed with PE (≥ 140/90 mmHg and/or the presence of a single incidence of proteinuria) [4], and participants with a normotensive pregnancy serving as the control group. All HIV positive women received antiretroviral therapy (ART).

#### Exclusion Criteria

women with polycystic ovarian syndrome, intrauterine death, cardiac disease, chorioamnionitis, unknown HIV status, eclampsia, sickle cell disease, active asthma that requires medication during the gestational period, abruption placentae, chronic renal disease, patients who have been declined from participation, systematic lupus erythematosus, pre-existing seizure disorders, and thyroid disease were not included in the study group.

### Multiplex Immunoassay

Maternal blood samples were collected and centrifuged at 3000 g for 10 minutes at 20°C. Serum was aliquoted and stored at -80°C until required. A cytometric mechanism with a bead-based flow constructed the assay, where multiplex analyses are permitted. Using the manufacturer’s instructions, Human Complement Magnetic Bead Panel (Millipore by Sigma Aldrich, catalogue number: HCMP1MAG-19K), Bio-Plex^®^MAGPIX™ (Bio Rad Laboratories, Inc., USA) was utilized, using the serum samples. Blank captured antibody with magnetic beads, C5a and C2 samples, antigen samples (1:4 dilution), and standards (serial dilution) were incubated overnight. Post incubation a triple wash eliminated any unbound substances. 50 µL of biotinylated detection antibody was added to each well. The assay plate was then incubated for 1 hour and washed to eliminate unbound biotinylated detection antibodies. Thereafter, 50 µL of 1X streptavidin-phycoerythrin (SA-PE) was added into each well. The plate was thereafter incubated for 10 minutes at 850 ± 50 rpm in a dark room. The assay plate was washed 3 times and re-suspended in assay buffer for 30 seconds at 850 ± 50rmp. Lastly, a Bio-Plex^®^MAGPIX™ Multiplex Reader (Bio Rad Laboratories, Inc., USA) was used to read the assay plate.

### Statistical Analysis

Data were analysed utilizing GraphPad Prism 5.00 for Windows (GraphPad Software, San Diego California USA). The Kolmogorov Smirnov normality test was used to check for parametric or non-parametric distribution. Non-parametric data are represented as median and interquartile range. Statistical significance according to pregnancy type (preeclamptic *vs* normotensive) and HIV status (negative *vs* positive) was determined using a Mann-Whitney’s *U* test. The Dunn’s Multiple Comparison *post hoc* test and the Kruskal-Wallis test determined statistical significance across all groups. A *p*-value of <0.05 was considered to be statistically significant. Spearman’s Rank Correlation Coefficient (*r*) was calculated to determine the relation between clinical/ demographic data versus C2 and C5a concentrations across the study population (-1 and 1). The outcome results were elucidated based on the degree of association as strong (0.7–1), moderate (0.5–0.7), or low (< 0.5) after taking significant correlation values into consideration.

## Results

### Patient Demographics and Clinical Characteristics

Patient demographics and clinical characteristics data are represented as median and interquartile range (IQR) in Table 1. Maternal age (*p =* 0.03), parity (*p* = 0.01), gravidity (*p* = 0.04), gestational age (*p* = 0.001), systolic (*p* < 0.0001) and diastolic blood pressures (*p* < 0.0001) were different across the study groups. However, maternal weight (*p* = 0.11) did not differ across all study groups.

**Table 1.** Patient demographics across study groups.

| **Total**<br>**Sample**<br>**Population**<br>**(N=76)** | **Normotensive**<br>**HIV Negative**<br>**(n=19)** | **Normotensive**<br>**HIV Positive**<br>**(n=19)** | **Preeclamptic**<br>**HIV Negative**<br>**(n=19)** | **Preeclamptic**<br>**HIV Positive**<br>**(n=19)** | ***p*-value** |
| --- | --- | --- | --- | --- | --- |
| **Weight**<br>**(kg)** | 74.00(22) | 81.00(28) | 90.00(46) | 79.50(35) | 0.1063 (ns) |
| **Gestational**<br>**Age**<br>**(weeks)** | 27.00 (9) | 25.00(14) | 24.00(10) | 23.00(10) | **0.0008\*\*\*** |
| **Parity** | 1.00(1) | 2.00(1) | 1.00(2) | 2.00(1) | **0.0085 \*** |
| **Systolic BP**<br>**(mmHg)** | 109.00(20) | 112.00(16) | 146.00(14) | 147.00(20) | **<0.0001 \*\*\*** |
| **Diastolic**<br>**BP**<br>**(mmHg)** | 65.00(13) | 72.00(14) | 92.00(9) | 97.00(13) | **<0.0001 \*\*\*** |
| **Maternal**<br>**Age (years)** | 25.00(9) | 31.00(11) | 29.00(16) | 34.00(14.50) | **0.0304\*** |
| **Gravidity** | 2.00(2) | 3.00(2) | 2.00(2) | 3.00(2) | **0.0425 \*** |

Results are represented as the median (IQR)
ns = non-significant,
\* $p < 0.05$
\*\* $p < 0.01$
\*\*\* $p < 0.001$

### Serum concentrations of complement component C2

#### Pregnancy type

C2 concentration was similar between the normotensive pregnant (median = 24285 pg/ml, IQR = 21319 pg/ml) *vs* the preeclamptic (median = 22287 pg/ml, IQR = 21701 pg/ml) groups, regardless of HIV status (Mann-Whitney U = 654; *p* = 0.4837; Fig 1A).

**Fig 1(A-C).**
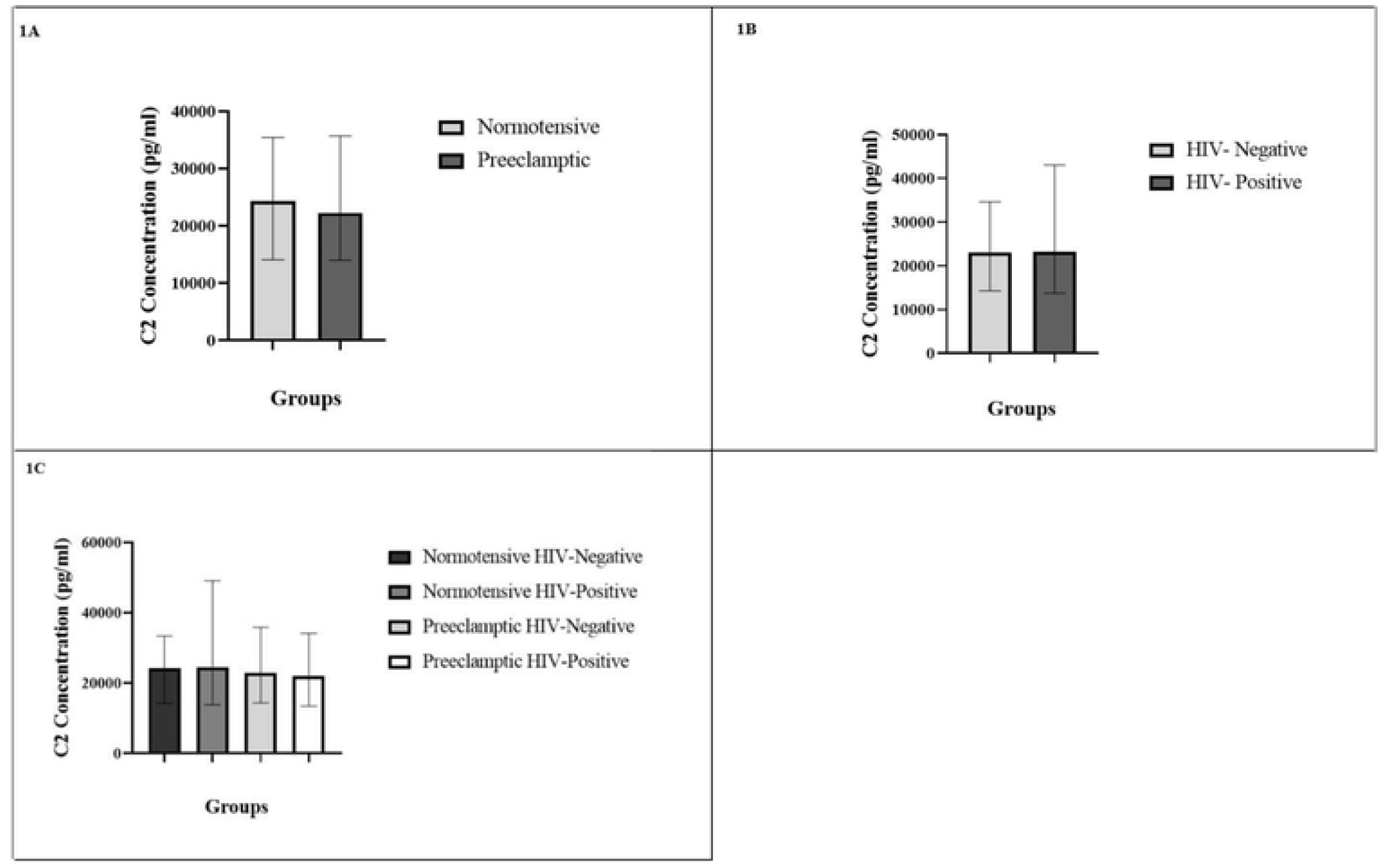
C2 concentration are depicted in: (A) Normotensive *vs* Preeclamptic groups, (B) HIV infected *vs* HIV uninfected groups and, (C) across all groups.

#### HIV status

The concentration of C2 level between the HIV positive (median = 23180 pg/ml, IQR = 29300 pg/ml) *vs* HIV negative (median = 23118 pg/ml, IQR = 20373 pg/ml) groups showed no statistical significance, irrespective of pregnancy type, (Mann-Whitney U = 690.5; *p* = 0.7469; Fig 1B).

#### Across all groups

The concentration of C2 was similar across all groups (Kruskal-Wallis H =1.098; *p* = 0.7776; Fig 2.3C; Table 2).

**Fig 2(A-E).**
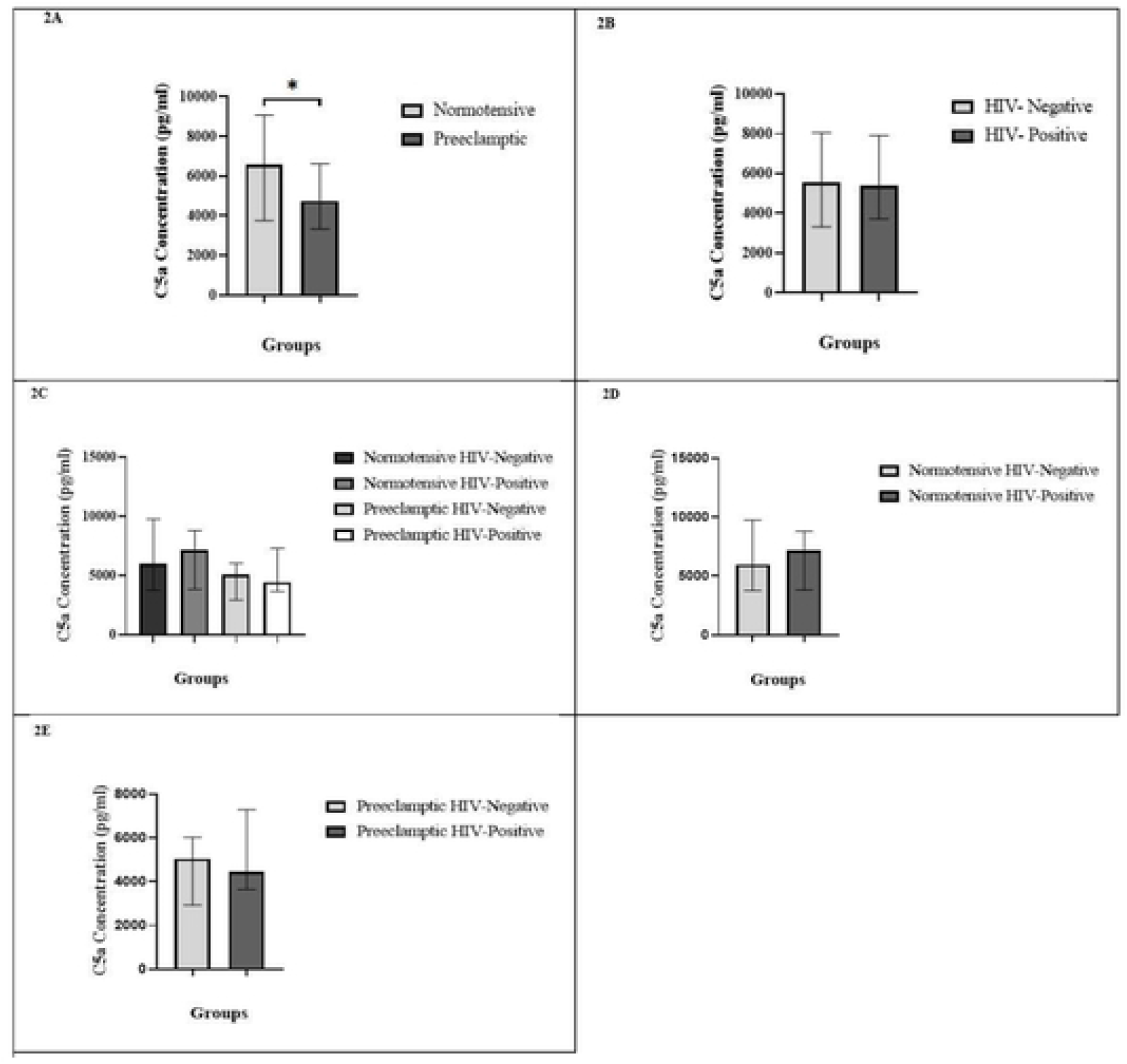
C5a concentration are depicted in: (A) Preeclamptic *vs* Normotensive groups, (B) HIV infected *vs* HIV uninfected groups, (C) across all groups. Serum concentration of C5a is significantly different between preeclamptic and normotensive groups, *p* = 0.0486. Serum concentration of C5a have no significant difference between HIV positive and HIV negative groups *p* = 0.8002, as well as across all groups *p* = 0.2259 (D) Normotensive HIV negative *vs* Normotensive HIV positive group and, (E) Preeclamptic HIV negative *vs* Preeclamptic HIV positive group.

**Fig 3.**
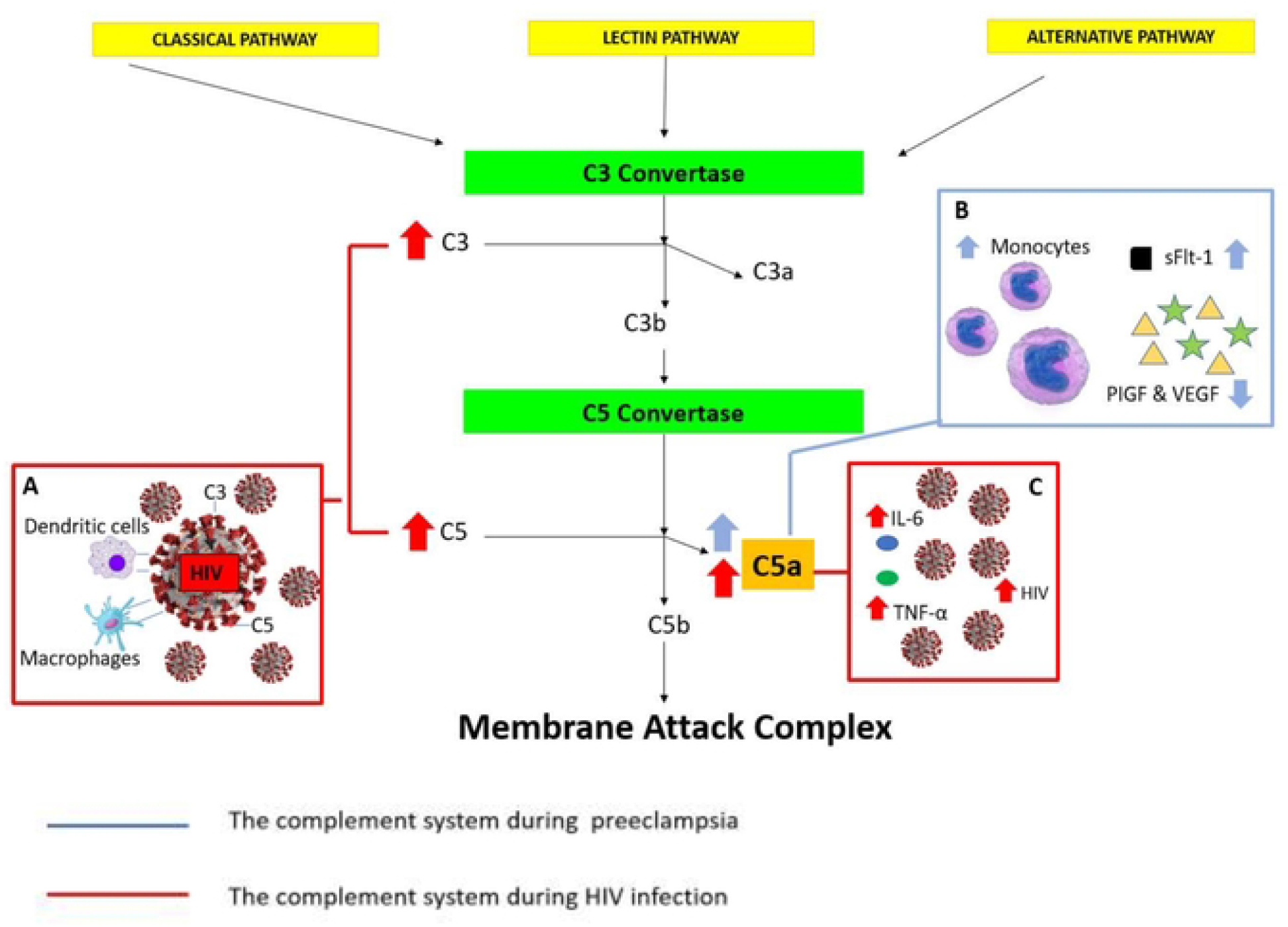
Schematic diagram showing the role of complement component C5a in the complement pathway of HIV associated preeclamptic women in **A:** Enhancement of C3 and C5 on the surface of HIV-1 enables the interaction of cells expressing complement receptors such as macrophages and dendritic cells (DC) with HIV-1 thereafter facilitating in HIV infection, **B:** Increased levels of C5a is associated with an increased number of monocytes. The escalation of C5a releases sFlt-1 resulting in the decrease of PIGF and VEGF, therefore, leading to preeclamptic development. **C:** C5a in excessive amounts is liable for the elevated production of pro-inflammatory cytokines, IL-6, and TNF-α and as a result, the promotion of HIV-1 infection and regulation.

**Table 2.** Serum concentration (pg/ml) of complement analytes across all study groups.

| Total Sample | Normotensive |  | Preeclamptic |  | p-value |
| --- | --- | --- | --- | --- | --- |
| Population<br>(N=76) | HIV Negative<br>(n=19) | HIV Positive<br>(n=19) | HIV Negative<br>(n=19) | HIV Positive<br>(n=19) |  |
| **C2**<br>(pg/ml) | 24185 (19167) | 24386 (35236) | 22929 (21506) | 22047 (20535) | 0.7776 |
| **C5a**<br>(pg/ml) | 5965 (6010) | 7160 (4958) | 5044 (3093) | 4447 (3642) | 0.2259 |

Results are represented as median (interquartile range)

### Serum concentrations of complement component C5a

#### Pregnancy type

The serum concentration of C5a was significantly higher in the normotensive pregnant (median = 6562 pg/ml, IQR = 5302 pg/ml) than the preeclamptic (median = 4745 pg/ml, IQR = 3279 pg/ml) group, regardless of HIV status (Mann-Whitney U = 532; *p* = 0.0486; Fig 2A).

#### HIV status

Serum C5a level was similar between the HIV positive (median = 5383 pg/ml, IQR = 4202 pg/ml) *vs* HIV negative (median = 5548 pg/ml, IQR = 4726 pg/ml) groups, irrespective of pregnancy type, (Mann-Whitney U = 697; *p* = 0.8002; Fig 2B).

#### Across all groups

The concentration of C5a did not differ across all groups (Kruskal-Wallis H =4.352; *p* = 0.2259; Fig 2C; Table 2).

#### Normotensive HIV status

No statistical significance was shown between the normotensive HIV negative group (median = 5965 pg/ml, IQR = 6010 pg/ml) *vs* normotensive HIV positive group (median = 7160 pg/ml, IQR = 4958 pg/ml). (Mann-Whitney U = 162; *p* = 0.6032; Fig 2D).

#### Preeclamptic HIV status

C5a expression was non-significantly different between the preeclamptic HIV negative group (median = 5044 pg/ml, IQR = 3093 pg/ml) *vs* preeclamptic HIV positive group (median = 4447 pg/ml, IQR = 3642 pg/ml); (Mann-Whitney U = 159; *p* = 0.5441; Fig 2E).

### Correlation between maternal demographics and serum C2 concentration

Diastolic blood pressure correlates with C2 concentration in normotensive HIV negative participants [*r* = - 0.463 (*p* < 0.05)] and in preeclamptic HIV negative participants [*r* = - 0.483 (*p* < 0.05)]. A negative correlation co-efficient demonstrated a relationship between maternal age and C2 [*r* = - 0.482 (*p* < 0.05)] in preeclamptic HIV positive patients. Table 4 displays the Spearman Rank Correlation Co-efficient for all other statistical non-significant maternal clinical/demographic data and C2 concentrations.

**Table 3.** Serum concentration (pg/ml) of C2 and C5a and Spearman Rank Correlation Co-Efficient (*r*) and level of significance (*p*).

| | C2<br>(pg/ml) | C5a<br>(pg/ml) | $r$ – value | $p$ – value |
| --- | --- | --- | --- | --- |
| **Normotensive** | 24285 (21319) | 6562 (5302) | 0.08856 | 0.5970 |
| **Preeclampsia** | 22287 (21701) | 4745 (3279) | 0.1534 | 0.3578 |
| **HIV Negative** | 22929 (20473) | 5548 (4726) | 0.2274 | 0.1759 |
| **HIV Positive** | 23180 (29300) | 5383 (4202) | 0.1089 | 0.5151 |

Results are represented as median (interquartile range)

**Table 4.**
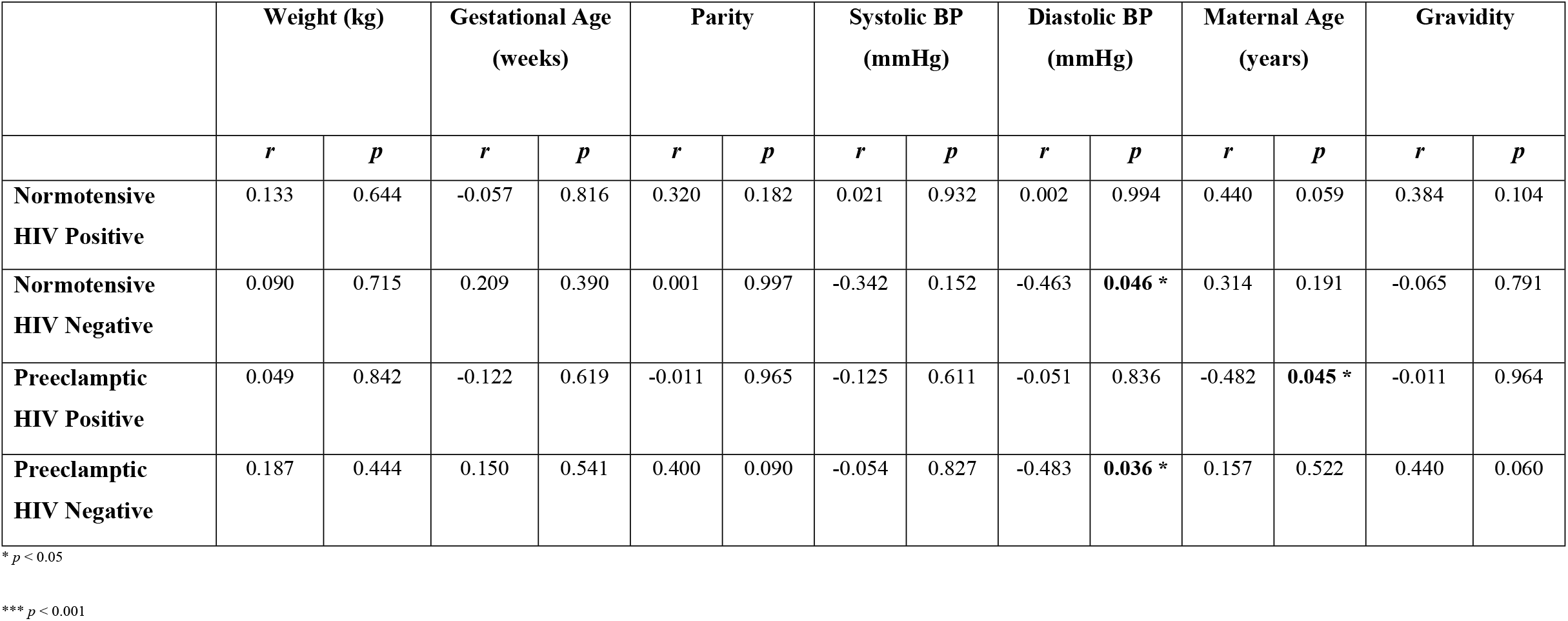
Serum concentration (pg/ml) of C2 and the Spearman’s correlation coefficient (*r*) and levels of significance (*p*)

|  | Weight (kg) |  | Gestational Age (weeks) |  | Parity |  | Systolic BP (mmHg) |  | Diastolic BP (mmHg) |  | Maternal Age (years) |  | Gravidity |  |
| --- | --- | --- | --- | --- | --- | --- | --- | --- | --- | --- | --- | --- | --- | --- |
|  | *r* | *p* | *r* | *p* | *r* | *p* | *r* | *p* | *r* | *p* | *r* | *p* | *r* | *p* |
| **Normotensive HIV Positive** | 0.133 | 0.644 | -0.057 | 0.816 | 0.320 | 0.182 | 0.021 | 0.932 | 0.002 | 0.994 | 0.440 | 0.059 | 0.384 | 0.104 |
| **Normotensive HIV Negative** | 0.090 | 0.715 | 0.209 | 0.390 | 0.001 | 0.997 | -0.342 | 0.152 | -0.463 | **0.046 \*** | 0.314 | 0.191 | -0.065 | 0.791 |
| **Preeclamptic HIV Positive** | 0.049 | 0.842 | -0.122 | 0.619 | -0.011 | 0.965 | -0.125 | 0.611 | -0.051 | 0.836 | -0.482 | **0.045 \*** | -0.011 | 0.964 |
| **Preeclamptic HIV Negative** | 0.187 | 0.444 | 0.150 | 0.541 | 0.400 | 0.090 | -0.054 | 0.827 | -0.483 | **0.036 \*** | 0.157 | 0.522 | 0.440 | 0.060 |

\* *p* < 0.05
\*\*\* *p* < 0.001

### Correlation between maternal demographics and serum C5a concentration

There was a significant correlation between gestational age and C5a concentration [*r* = -0.523 (*p* < 0.05)] in normotensive HIV positive patients. Additionally, a positive correlation co-efficient *r*= 0.615 (*p* < 0.001) was noted between diastolic BP and C5a concentration preeclamptic HIV positive group, likewise a positive correlation was observed between systolic blood pressure and C5a concentration *r* = 0.483 (*p* < 0.05) in preeclamptic HIV negative patients (Table 5).

**Table 5.** Serum concentration (pg/ml) of C5a and the Spearman’s correlation coefficient (*r*) and levels of significance (*p*)

|  | Weight (kg) |  | Gestational Age (weeks) |  | Parity |  | Systolic BP (mmHg) |  | Diastolic BP (mmHg) |  | Maternal Age (years) |  | Gravidity |  |
| --- | --- | --- | --- | --- | --- | --- | --- | --- | --- | --- | --- | --- | --- | --- |
|  | *r* | *p* | *r* | *p* | *r* | *p* | *r* | *p* | *r* | *p* | *r* | *p* | *r* | *p* |
| **Normotensive HIV Positive** | 0.442 | 0.058 | -0.523 | 0.022 * | 0.163 | 0.505 | -0.182 | 0.455 | 0.023 | 0.926 | 0.165 | 0.498 | 0.125 | 0.610 |
| **Normotensive HIV Negative** | 0.206 | 0.397 | 0.318 | 0.185 | 0.341 | 0.154 | -0.104 | 0.670 | -0.447 | 0.055 | 0.362 | 0.127 | -0.346 | 0.147 |
| **Preeclamptic HIV Positive** | -0.016 | 0.949 | -0.027 | 0.912 | -0.050 | 0.839 | 0.276 | 0.253 | 0.615 | **0.005 \*\*\*** | -0.260 | 0.297 | 0.047 | 0.849 |
| **Preeclamptic HIV Negative** | -0.159 | 0.516 | 0.291 | 0.227 | -0.134 | 0.584 | 0.483 | **0.036 \*** | -0.344 | 0.149 | -0.155 | 0.526 | -0.048 | 0.847 |

\* *p* < 0.05
\*\*\* *p* < 0.001

### Relationship of C2 and C5a

There was no statistically significant correlation of C2 on C5a concentration and vice versa (Table 4).

## Discussion

Our study demonstrates a down-regulation of serum C5a concentration in PE compared to normotensive pregnancies, irrespective of HIV status at term. This unexpected finding may be attributed to C5a being consumed faster than production, resulting in its rapid removal from circulation [32]. Alternatively, there may be a dysregulation of its receptor (C5aR) or ligand-receptor mediated affinity thereby inhibiting signal transduction. Off note, a knockdown of C5aR with small interfering RNA (siRNA) salvages endothelial cell migration and vessel formation [33]. Endothelial injury and angiogenic inhibition characterises PE development. In normal pregnancy, there is a mild systemic inflammation in response to the semi-allogenic fetus whilst in PE there is excessive maternal inflammation [34]. Notably, the anaphylatoxin C5a represents fragments of activated complement proteins that are the main mediators of an inflammatory response. A study conducted by Burwick *et al*. (2013) demonstrated that plasma concentrations of C5a and C5b-9 are exaggerated in PE, this leads to the urinary excretion of excess C3a, C5a, and C5b-9 [32]. Richani *et al*. (2005) showed that plasma C5a concentration is elevated in normotensive pregnancies compared to non-pregnant women respectively [16, 35]. These results may be due to inadequate renal clearance/filtration of these circulating complement proteins via [36]. This suggests that complement dysregulation in ongoing disease arises mainly at the level of C5 [32] and may be a possible explanation for the downregulation of C5a during gestation as observed in PE in our study.

Notably, C5a exerts its damaging effect by inducing the release of the potent anti-angiogenic factor, soluble vascular endothelial growth factor receptor-1 (sVEGFR-1), also known as sFlt-1 [26] (Fig 3). It is widely accepted that PE is associated with a rise of this scavenger receptor, sFlt-1. This elevation with a concurrent decline in VEGF and PlGF inhibits endothelial cell migration and tube formation [26]. Moreover, Govender *et al*. (2013) also reported elevated sEng in PE, regardless of HIV infection supporting an offset of the immune hyperactivity [9]. sEng weakens TGF-β1 receptor transduction and prevents activation of the endothelial nitric oxide synthase 3 (eNOS) action, thereby promoting the development of hypertension. The C5a downregulation in our study, is confounding and may be attributed to our small sample size and ART. In a study by Burwick and Feinberg (2013), Eculizumab a monoclonal antibody inhibitor of C5 decreases the production of complement components C5a and C5b-9 and their downstream effects [37,38]. Worldwide, C5 deficiency is correlated with genetic defects. It is possible that a polymorphism of the C5 gene may be involved in C5a dysregulation, in fact, a complement C5 gene, c.754G>A:p.A252T mutation has been demonstrated in the Western Cape, South Africa in Black African patients infected with meningococcal disease [39-41]. A single nucleotide polymorphism in maternal C5, C5a and fetal CD55 and CD59 may constitute independent risk factors for PE development. It would be interesting to examine the C5a gene in Black African women of isiZulu origin in South Africa.

Our study also reports similar C5a levels between HIV negative compared to HIV positive women, regardless of pregnancy type. It is widely accepted that this pattern recognition component is activated during HIV infection because C5a serves to attract dendritic cells and macrophages to sites of HIV entry [42]. Of note, C5aR1 is critical for viral entry [43]. In fact, the targeted reduction of C5aR1 expression reduces HIV infection by 50% because C5aR1 acts as an enhancer of CCR5-mediated HIV entry into macrophages (Fig 3). It is therefore plausible that the similar C5a levels may be attributed to a dysregulation of C5, C5aR1 expression, or ART usage. Nonetheless, C5a has a small amphipathic configuration with an antiviral action [44, 45] that blocks herpes simplex virus 1 (HSV-1), HSV-2, hepatitis C, and HIV-1 entry by disrupting the integrity of viral membranes [46-48]. Also, HIV induces the cleavage of C5 to generate the anaphylatoxin C5a, which attracts immature dendritic cells (DC) to promote viral amplification and dissemination [42]. A plausible explanation for the similarity between HIV negative and HIV positive groups in our study may be the inadequate binding of C5a to C5aR, mediated by the effect of ART [42]. More specifically, the removal of C-terminal arginyl residues from C5a by the enzyme carboxypeptidase N converts it into a proinflammatory form called Desarginated C5a (C5a^desArg^) [49]; a potent stimulatory factor that primes monocyte-derived macrophages for HIV entry. Furthermore, kinetic analyses of HIV replication have shown that exposure to C5a leads to the acceleration of HIV infection in macrophages [25]. Moreover, C5a independently induces release of pro-inflammatory Tumour Necrosis Factor-alpha (TNF-α), IL-1, and Interleukin 6 (IL-6) by monocytes and macrophages [50, 51]. This increase of pro-inflammatory cytokines TNF-α and IL-6 occur in the presence of C5a, and both are known promoters of HIV-1 infection and regulation [25,28]. Maharaj *et al*. (2017) reported a decrease of IL-2 and TNF-α concentrations in PE and a lower IFN-γ and IL-6 concentrations in HIV-infected preeclamptics receiving HAART relative to uninfected preeclamptic women. HIV infection together with HAART lowers the production of inflammatory cytokines during pregnancy in both successful and preeclamptic pregnancies [52]. It is plausible to hypothesize that C5a production has a directly proportional effect on the release of cytokine TNF-α and IL-6. Since these cytokines are reduced in HIV-infected patients, one may deduce that decline emanates from the lower C5a expression.

In addition, we report similar C2 expression between PE and normotensive pregnancies, irrespective of HIV status albeit a down regulatory trend. This observation in PE is consistent with previous reports where C2 polymorphisms and its deficiency is linked to chronic inflammatory conditions such as systemic lupus erymatosus (SLE) [53, 54]. The clinical manifestations of PE mimics the exacerbated immune microenvironment of SLE, where a 10% penetrance of C2 deficiency occurs [30]. Moreover, in pregnant women with SLE, there is complement-mediated injury, predisposing them to a greater risk of PE development, placental insufficiency, miscarriage, and fetal growth restriction [55]. It is widely accepted that the complement cascade is activated in PE via the LP and/or the CP [56]. Activation of these pathways are impaired in C2 deficient patients where during periods of active disease, serum complement activity is reduced emanating from a low expression of CP components notably C1q, C2, C4 (Fig 3) [57]. Also, C2 deficiency predominates in females [58]. Hepatocytes synthesize 90% of plasma complement components [59]. Since liver enzyme abnormalities occur in approximately 10% of PE [60], it is plausible that the C2 and C5a downregulation in our study may be attributed to liver dysfunction in the PE cohort. In fact, in C5 deficient mice, the administration of murine C5 or C5a restores hepatocyte regeneration while obstruction of C5aR prevents hepatocyte proliferation [61].

We also demonstrate that HIV status did not affect serum C2 expression at term, albeit with an upregulated trend. These results are similar to that of Mahajan *et al*. (2017) who reported *in vitro* studies of HIV-1 infected astrocytic cell lines and primary astrocytes a significant enhancement of C2 and C3 expression [62]. Huson *et al*. (2015) reported increased C3 and C1q-C4 levels in asymptomatic patients with HIV infection compared to healthy controls [31]. The similar C2 expression in our study may be attributed to HAART the standard treatment for all HIV positive patients in South Africa (SA). It is plausible that HAART lowers C2 levels to re-instate immune response [63]. In an astrocytic U373 cell line, HIV-1 increases mRNA level of C2 [64]. Moreover, the regulation of C2 production may be attributed to HIV viral proteins Nef, Rev, and Tat on the complement promoter protein synthesis in the host cell. Furthermore, structural proteins such as gp120 and gp41 may also regulate HIV expression [65]. Furthermore, the modulation of C2 production may be secondary emanating from HIV-induced transcription factor NFκB, the main controlling factor in viral transcription.

In our study, we report a moderate correlation between gestational age (*r*=-0.523), diastolic (*r*=0.615) and systolic blood pressure (*r*=0.483) with C5a concentration in normotensive HIV positive, preeclamptic HIV positive and preeclamptic HIV negative respectively. High blood pressure has been previously correlated with elevated C5a in humans [66]. Notably, the development of hypertension is also associated with the development of vascular damage [67, 68]. In our study, gestational age was negatively correlated with C5a concentration [*r* = -0.523 (*p* < 0.05)] in normotensive HIV positive group. Whilst normal pregnancy is associated with complement activation, gestational age does not correlate with the anaphylatoxin C5a [16]. We also report a moderate negative correlation between diastolic blood pressure and C2 concentration in the normotensive HIV negative [*r* = - 0.463 (*p* < 0.05)] and preeclamptic HIV negative [*r* = - 0.483 (*p* < 0.05)] participants. In hypertension, C3, C4 and C5 levels, all by-products of C2 correlate with development of hypertension [67]. C2 downregulation has been previously linked with age-related macular degeneration and genetic polymorphisms in people of Indian ethnicity [69]. Maternal age is a predisposing factor to PE development [70]. This study also reports no significant correlation between C2 concentration on C5a and vice versa. Limitations of our study that may have influenced analyte expression include the small sample size, all HIV infected women received ART and the unknown duration of ART/HAART management.

## Conclusion

This novel study reports a significant downregulation of C5a in PE. This observation suggests a loss of regulation of complement activation, dysregulation of signal transduction and/ or polymorphisms of C5a in women of African descent. We also demonstrate a similar C5a and C2 expression by HIV status, which may be attributed to ART/HAART therapy. Notably, the dysregulation of complement activation will impact the host innate defence, enhancing placental and fetal injury. Further studies should include a large cohort that investigates C2, C5a, C5a-desArg, C5aR1 and C5aR2 together with pro-inflammatory polypeptides in the development of PE. Single nucleotide polymorphisms investigations of these components are required to better understand the complement cascade in complicated pregnancies thereby preventing premature delivery and enabling better clinical management in the synergy of HIV and preeclampsia.

## Data Availability

N/A

## Acknowledgments

The authors wish to thank Wided Kelmemi for technical assistance.

## Financial statement

The laboratory expense for this project was covered by the UKZN productivity award of Professor T Naicker.

## References

1. Girum, T. & Wasie, A. (2017). Correlates of maternal mortality in developing countries: an ecological study in 82 countries. Maternal Health, Neonatology and Perinatology, 3(1): 19.

2. National Committee for Confidential Enquiry into Maternal Deaths. Saving mothers Report. (2017). Annual report on the Confidential Enquiries into Maternal Deaths in South Africa. Pretoria: South African Department of Health, 2019.

3. Mdlalose, S., Moodley, J. & Naicker, T. (2020). The role of follistatin and granulocyte-colony stimulating factor in HIV-associated pre-eclampsia. Pregnancy Hypertension, 19(1): 81–86.

4. Brown, M. A., Magee, L. A., Kenny, L. C., Karumanchi, S. A., Mccarthy, F. P., Saito, S., Hall, D. R., Warren, C. E., Adoyi, G. & Ishaku, S. (2018). Hypertensive disorders of pregnancy: ISSHP classification, diagnosis, and management recommendations for international practice. Hypertension, 72(1): 24–43.

5. Young, B. C., Levine, R. J. & Karumanchi, S. A. (2010). Pathogenesis of Preeclampsia. Review of Pathology: Mechanisms of Disease, 5(1): 173–92.

6. Regal, J. F., Burwick, R. M. & Fleming, S. D. (2017). The Complement System and Preeclampsia. Current Hypertension Reports, 19(11): 87.

7. Silasi, M., Cohen, B., Karumanchi, S. A. & Rana, S. (2010). Abnormal placentation, angiogenic factors, and the pathogenesis of preeclampsia. Obstetrics and Gynecology Clinics, 37(2): 239–253.

8. Hsu, P. & Nanan, R. K. H. (2014). Innate and adaptive immune interactions at the fetal– maternal interface in healthy human pregnancy and pre-eclampsia. Frontiers in Immunology, 5(1): 125.

9. Govender, N., Naicker, T. & Moodley, J. (2013). Maternal imbalance between pro-angiogenic and anti-angiogenic factors in HIV-infected women with pre-eclampsia. Cardiovascular Journal of Africa, 24(5): 174.

10. Hall, D., Gebhardt, S., Theron, G. & Grové, D. (2014). Pre-eclampsia and gestational hypertension are less common in HIV infected women. Pregnancy Hypertension: An International Journal of Women’s Cardiovascular Health, 4(1): 91–96.

11. Kalumba, V., Moodley, J. & Naidoo, T. (2013). Is the prevalence of pre-eclampsia affected by HIV/AIDS? A retrospective case–control study. Cardiovascular Journal of Africa, 24(2): 24.

12. Phoswa, W. N., Naicker, T., Ramsuran, V. & Moodley, J. (2019). Pre-eclampsia: the role of highly active antiretroviral therapy and immune markers. Inflammation Research, 68(1): 47–57.

13. Fakhouri, F. (2016). Pregnancy-related thrombotic microangiopathies: clues from complement biology. Transfusion and Apheresis Science, 54(2): 199–202.

14. Sabau, L., Terriou, L., Provot, F., Fourrier, F., Roumier, C., Caron, C., Susen, S. & Ducloy-Bouthors, A. (2016). Are there any additional mechanisms for haemolysis in HELLP syndrome? Thrombosis Research, 142(1): 40–43.

15. Alrahmani, L. & Willrich, M. A. V. (2018). The complement alternative pathway and preeclampsia. Current Hypertension Reports, 20(5): 40.

16. Richani, K., Soto, E., Romero, R., Espinoza, J., Chaiworapongsa, T., Nien, J. K., Edwin, S., Kim, Y. M., Hong, J.-S. & Mazor, M. (2005). Normal pregnancy is characterized by systemic activation of the complement system. The Journal of Maternal-Fetal & Neonatal Medicine, 17(4): 239–245.

17. Khan, R., Maduray, K., Moodley, J. & Naicker, T. (2016). Activation of CD35 and CD55 in HIV associated normal and pre-eclamptic pregnant women. European Journal of Obstetrics & Gynecology and Reproductive Biology, 100(204): 51–56.

18. Merle, N. S., Church, S. E., Fremeaux-Bacchi, V. & Roumenina, L. T. (2015). Complement system part I–molecular mechanisms of activation and regulation. Frontiers in Immunology, 6(1): 262.

19. Wills-Karp, M. (2007). Complement activation pathways: a bridge between innate and adaptive immune responses in asthma. Proceedings of the American Thoracic Society, 4(3): 247–251.

20. Lillegard, K. E., Johnson, A. C., Lojovich, S. J., Bauer, A. J., Marsh, H. C., Gilbert, J. S. & Regal, J. F. (2013). Complement activation is critical for placental ischemia-induced hypertension in the rat. Molecular Immunology, 56(1-2): 91–97.

21. Mcdonald, C. R., Tran, V. & Kain, K. C. (2015). Complement activation in placental malaria. Frontiers in Microbiology, 6(1): 1460.

22. Yu, Q., Yu, R. & Qin, X. (2010). The good and evil of complement activation in HIV-1 infection. Cellular & Molecular Immunology, 7(5): 334–340.

23. Klos, A., Tenner, A. J., Johswich, K. O., Ager, R. R., Reis, E. S. & Köhl, J. (2009). The role of the anaphylatoxins in health and disease. Molecular Immunology, 46(14): 2753–66.

24. Sarma, J. V. & Ward, P. A. (2011). The complement system. Cell and Tissue Research, 343(1): 227–235.

25. Kacani, L., Bánki, Z., Zwirner, J., Schennach, H., Bajtay, Z., Erdei, A., Stoiber, H. & Dierich, M. P. (2001). C5a and C5adesArg enhance the susceptibility of monocyte-derived macrophages to HIV infection. The Journal of Immunology, 166(5): 3410–3415.

26. Girardi, G., Prohászka, Z., Bulla, R., Tedesco, F. & Scherjon, S. (2011). Complement activation in animal and human pregnancies as a model for immunological recognition. Molecular Immunology, 48(14): 1621–1630.

27. Conroy, A., Serghides, L., Finney, C., Owino, S. O., Kumar, S., Gowda, D. C., Liles, W. C., Moore, J. M. & Kain, K. C. (2009). C5a enhances dysregulated inflammatory and angiogenic responses to malaria in vitro: potential implications for placental malaria. PloS One, 4(3): e4953.

28. Popko, K., Gorska, E., Stelmaszczyk-Emmel, A., Plywaczewski, R., Stoklosa, A., Gorecka, D., Pyrzak, B. & Demkow, U. (2010). Proinflammatory cytokines Il-6 and TNF-α and the development of inflammation in obese subjects. European Journal of Medical Research, 15(S2): 120.

29. Morris Jr, J. A., Francois, C., Olson, P. K., Cotton, B. A., Summar, M., Jenkins, J. M., Norris, P. R., Moore, J. H., Williams, A. E. & Mcnew, B. S. (2009). Genetic variation in complement component 2 of the classical complement pathway is associated with increased mortality and infection: a study of 627 trauma patients. The Journal of Trauma, 66(5): 1265.

30. Lintner, K. E., Wu, Y. L., Yang, Y., Spencer, C. H., Hauptmann, G., Hebert, L. A., Atkinson, J. & Yu, C. Y. (2016). Early components of the complement classical activation pathway in human systemic autoimmune diseases. Frontiers in Immunology, 7(FEB): 36.

31. Huson, M. A., Wouters, D., Van Mierlo, G., Grobusch, M. P., Zeerleder, S. S. & Van Der Poll, T. (2015). HIV coinfection enhances complement activation during sepsis. The Journal of Infectious Diseases, 212(3): 474–483.

32. Burwick, R. M., Fichorova, R. N., Dawood, H. Y., Yamamoto, H. S. & Feinberg, B. B. (2013). Urinary excretion of C5b-9 in severe preeclampsia: tipping the balance of complement activation in pregnancy. Hypertension, 62(6): 1040–1045.

33. Ma, Y., Kong, L. R., Ge, Q., Lu, Y. Y., Hong, M. N., Zhang, Y., Ruan, C. C. & Gao, P. J. (2018). Complement 5a-mediated trophoblasts dysfunction is involved in the development of pre-eclampsia. Journal of Cellular and Molecular Medicine, 22(2): 1034–1046.

34. Chaouat, G., Sandra, O. & Lédée, N. (2013). Immunology of Pregnancy 2013, Bentham Science Publishers.

35. Denny, K. J., Coulthard, L. G., Finnell, R. H., Callaway, L. K., Taylor, S. M. & Woodruff, T. M. (2013). Elevated complement factor C5a in maternal and umbilical cord plasma in preeclampsia. Journal of Reproductive Immunology, 97(2): 211–216.

36. Oppermann, M. & Götze, O. (1994). Plasma clearance of the human C5a anaphylatoxin by binding to leucocyte C5a receptors. Immunology, 82(4): 516.

37. Burwick, R. & Feinberg, B. (2013). Eculizumab for the treatment of preeclampsia/HELLP syndrome. Placenta, 34(2): 201–203.

38. Thomas, T. C., Rollins, S. A., Rother, R. P., Giannoni, M. A., Hartman, S. L., Elliott, E. A., Nye, S. H., Matis, L. A., Squinto, S. P. & Evans, M. J. (1996). Inhibition of complement activity by humanized anti-C5 antibody and single-chain Fv. Molecular Immunology, 33(17-18): 1389–1401.

39. Aguilar-Ramirez, P., Reis, E., Florido, M., Barbosa, A., Farah, C., Costa-Carvalho, B. & Isaac, L. (2009). Skipping of exon 30 in C5 gene results in complete human C5 deficiency and demonstrates the importance of C5d and CUB domains for stability. Molecular Immunology, 46(10): 2116–2123.

40. López-Lera, A., Garrido, S., De La Cruz, R. M., Fontán, G. & López-Trascasa, M. (2009). Molecular characterization of three new mutations causing C5 deficiency in two non-related families. Molecular Immunology, 46(11-12): 2340–2347.

41. Schejbel, L., Fadnes, D., Permin, H., Lappegård, K. T., Garred, P. & Mollnes, T. E. (2013). Primary complement C5 deficiencies–molecular characterization and clinical review of two families. Immunobiology, 218(10): 1304–1310.

42. Soederholm, A., Bánki, Z., Wilflingseder, D., Gassner, C., Zwirner, J., López-Trascasa, M., Falkensammer, B., Dierich, M. P. & Stoiber, H. (2007). HIV-1 induced generation of C5a attracts immature dendriticcells and promotes infection of autologous T cells. European Journal of Immunology, 37(8): 2156–2163.

43. Moreno-Fernandez, M. E., Aliberti, J., Groeneweg, S., Köhl, J. & Chougnet, C. A. (2016). A novel role for the receptor of the complement cleavage fragment C5a, C5aR1, in CCR5-mediated entry of HIV into macrophages. AIDS Research and Human Retroviruses, 32(4): 399–408.

44. Cheng, G., Montero, A., Gastaminza, P., Whitten-Bauer, C., Wieland, S. F., Isogawa, M., Fredericksen, B., Selvarajah, S., Gallay, P. A. & Ghadiri, M. R. (2008). A virocidal amphipathic α-helical peptide that inhibits hepatitis C virus infection in vitro. Proceedings of the National Academy of Sciences, 105(8): 3088–3093.

45. Veazey, R. S., Chatterji, U., Bobardt, M., Russell-Lodrigue, K. E., Li, J., Wang, X. & Gallay, P. A. (2016). C5A protects macaques from vaginal simian-human immunodeficiency virus challenge. Antimicrobial Agents and Chemotherapy, 60(1): 693–698.

46. Brass, V., Bieck, E., Montserret, R., Wölk, B., Hellings, J. A., Blum, H. E., Penin, F. & Moradpour, D. (2002). An amino-terminal amphipathic α-helix mediates membrane association of the hepatitis C virus nonstructural protein 5A. Journal of Biological Chemistry, 277(10): 8130–8139.

47. Stupar, R. M. (2010). Into the wild: The soybean genome meets its undomesticated relative. Proceedings of the National Academy of Sciences, 107(51): 21947–21948.

48. De Witte, L., Bobardt, M. D., Chatterji, U., Van Loenen, F. B., Verjans, G. M., Geijtenbeek, T. B. & Gallay, P. A. (2011). HSV neutralization by the microbicidal candidate C5A. PloS One, 6(5): e18917.

49. Zwirner, J., Götze, O., Sieber, A., Kapp, A., Begemann, G., Zuberbier, T. & Werfel, T. (1998). The human mast cell line HMC-1 binds and responds to C3a but not C3a (desArg). Scandinavian Journal of Immunology, 47(1): 19–24.

50. Cavaillon, J. M., Fitting, C. & Haeffner-Cavaillon, N. (1990). Recombinant C5a enhances interleukin 1 and tumor necrosis factor release by lipopolysaccharide-stimulated monocytes and macrophages. European Journal of Immunology, 20(2): 253–257.

51. Montz, H., Koch, K., Zierz, R. & Götze, O. (1991). The role of C5a in interleukin-6 production induced by lipopolysaccharide or interleukin-1. Immunology, 74(3): 373.

52. Maharaj, N. R., Phulukdaree, A., Nagiah, S., Ramkaran, P., Tiloke, C. & Chuturgoon, A. A. (2017). Pro-inflammatory cytokine levels in HIV infected and uninfected pregnant women with and without preeclampsia. PLoS One, 12(1): e0170063.

53. Agnello, V. (1978). Association of systemic lupus erythematosus and SLE-like syndromes with hereditary and acquired complement deficiency states. Arthritis and Rheumatism, 21(S5): S146–52.

54. Macedo, A. C. L. & Isaac, L. (2016). Systemic lupus erythematosus and deficiencies of early components of the complement classical pathway. Frontiers in Immunology, 7(1): 55.

55. Teirilä, L., Heikkinen-Eloranta, J., Kotimaa, J., Meri, S. & Lokki, A. I. (2019). Regulation of the complement system and immunological tolerance in pregnancy. Complement Regulation, 45(1): 18.

56. Derzsy, Z., Prohászka, Z., Rigó Jr, J., Füst, G. & Molvarec, A. (2010). Activation of the complement system in normal pregnancy and preeclampsia. Molecular Immunology, 47(7-8): 1500–1506.

57. Pickering, M. & Walport, M. (2000). Links between complement abnormalities and systemic lupus erythematosus. Rheumatology, 39(2): 133–141.

58. Lynch, A. M. & Salmon, J. E. (2010). Dysregulated complement activation as a common pathway of injury in preeclampsia and other pregnancy complications. Placenta, 31(7): 561–567.

59. Morgan, B. & Gasque, P. (1997). Extrahepatic complement biosynthesis: where, when and why?. Clinical and Experimental Immunology, 107(1): 1.

60. Knox, T. A. & Olans, L. B. (1996). Liver disease in pregnancy. New England Journal of Medicine, 335(8): 569–576.

61. Qin, X. & Gao, B. (2006). The complement system in liver diseases. Cellular & Molecular Immunology, 3(5): 333–340.

62. Mahajan, S. D., Aalinkeel, R., Parikh, N. U., Jacob, A., Cwiklinski, K., Sandhu, P., Le, K., Loftus, A. W., Schwartz, S. A. & Quigg, R. J. (2017). Immunomodulatory role of complement proteins in the neuropathology associated with opiate abuse and HIV-1 Co-morbidity. Immunological Investigations, 46(8): 816–832.

63. Pillay, Y., Moodley, J. & Naicker, T. (2019). The role of the complement system in HIV infection and preeclampsia. Inflammation Research, 68(6): 459–469.

64. Speth, C., Stöckl, G., Mohsenipour, I., Würzner, R., Stoiber, H., Lass-Flörl, C. & Dierich, M. P. (2001). Human immunodeficiency virus type 1 induces expression of complement factors in human astrocytes. Journal of Virology, 75(6): 2604–2615.

65. Corasaniti, M. T., Bagetta, G., Rotiroti, D. & Nisticò, G. (1998). The HIV envelope protein gp120 in the nervous system: interactions with nitric oxide, interleukin-1β and nerve growth factor signalling, with pathological implications in vivo and in vitro. Biochemical Pharmacology, 56(2): 153–156.

66. Zhang, C., Li, Y., Wang, C., Wu, Y., Cui, W., Miwa, T., Sato, S., Li, H., Song, W.-C. & Du, J. (2014). Complement 5a receptor mediates angiotensin II–induced cardiac inflammation and remodeling. Arteriosclerosis, Thrombosis, and Vascular Biology, 34(6): 1240–1248.

67. Ruan, C.-C. & Gao, P.-J. (2019). Role of complement-related inflammation and vascular dysfunction in hypertension. Hypertension, 73(5): 965–971.

68. Wenzel, U. O., Kemper, C. & Bode, M. (2020). The role of complement in arterial hypertension and hypertensive end organ damage. British Journal of Pharmacology.1(9): 271

69. Kaur, I., Katta, S., Reddy, R. K., Narayanan, R., Mathai, A., Majji, A. B. & Chakrabarti, S. (2010). The involvement of complement factor B and complement component C2 in an Indian cohort with age-related macular degeneration. Investigative Ophthalmology & Visual Science, 51(1): 59–63.

70. Whitelaw, N., Bhattacharya, S., Mclernon, D. & Black, M. (2014). Internet information on birth options after caesarean compared to the RCOG patient information leaflet; a web survey. BMC Pregnancy and Childbirth, 14(1): 1–13.

